# Minimal observed impact of HLA genotype on hospitalization and severity of SARS-CoV-2 infection

**DOI:** 10.1101/2021.12.22.21268062

**Authors:** Austin Nguyen, Tasneem Yusufali, Jill A. Hollenbach, Abhinav Nellore, Reid F. Thompson

**Affiliations:** Department of Biomedical Engineering; Oregon Health & Science University; Portland, OR, 97239; USA; Department of Neurology; University of California, San Francisco; San Francisco, CA, 94158; USA; Department of Epidemiology and Biostatistics; University of California, San Francisco; San Francisco, CA, 94158; USA; Department of Surgery; Oregon Health & Science University; Portland, OR, 97239; USA; Department of Radiation Medicine; Oregon Health & Science University; Portland, OR, 97239; USA; Department of Medical Informatics and Clinical Epidemiology; Oregon Health & Science University; Portland, OR, 97239; USA; Division of Hospital and Specialty Medicine; VA Portland Healthcare System; Portland, OR, 97239; USA

## Abstract

HLA is a critical component of the viral antigen presentation pathway. We investigated the relationship between severity of SARS-CoV-2 disease and HLA type in 3,235 individuals with confirmed SARS-CoV-2 infection. We found only the DPB1 locus to be associated with the binary outcome of whether an individual developed any COVID-19 symptoms. The number of peptides predicted to bind to an HLA allele had no significant relationship with disease severity both when stratifying individuals by ancestry or age and in a pooled analysis. Age, BMI, asthma status, and autoimmune disorder status were predictive of severity across multiple age and individual ancestry stratificiations. Overall, at the population level, we found HLA type is significantly less predictive of COVID-19 disease severity than certain demographic factors and clinical comorbidities.

## BACKGROUND

The global COVID-19 pandemic has exposed significant gaps in our ability to predict disease trajectory among individuals, with many people experiencing asymptomatic infections while others may be hospitalized with or die from COVID-19. Observational analyses have identified disease severity risk factors such as age, BMI, and sex (1–3). However, host immunogenetic factors such as human leukocyte antigen (HLA) type may help determine the severity of SARS-CoV-2 infection. HLA is a critical component of the viral antigen presentation pathway, and previous studies have shown that individual HLA alleles may confer differential susceptibility and severity across viral diseases, including SARS-CoV-2 (4–11).

While large genotype association studies have investigated the relationship between genetic variants and severity of COVID-19 disease, they have not generally implicated the HLA locus (10,12–14). Further, while a growing collection of single institution or regional hospital system-based studies have reported HLA associations with COVID-19 disease (15–18), the statistical significance of these associations does not withstand correction for multiple comparisons. The relationship between HLA genotype and severity of COVID-19 disease, especially across a large and diverse population, thus remains unclear. In this study, we investigated the specific relationship between HLA type and COVID-19 severity in a cohort of 3,235 individuals obtained from AncestryDNA (10,19) with confirmed SARS-CoV-2 infection.

## RESULTS

We extracted basic demographic and clinical data for 3,235 individuals among the AncestryDNA cohort (10,19) with a positive SARS-CoV-2 nasal swab and classified the severity of their COVID-19 disease according to patient survey responses (Table 1). We next assessed the extent to which these demographic and clinical features predicted COVID-19 severity, and we found comorbidities that are contributors in a linear model predicting hospitalization (Supplementary Table 1).

**Table 1:**
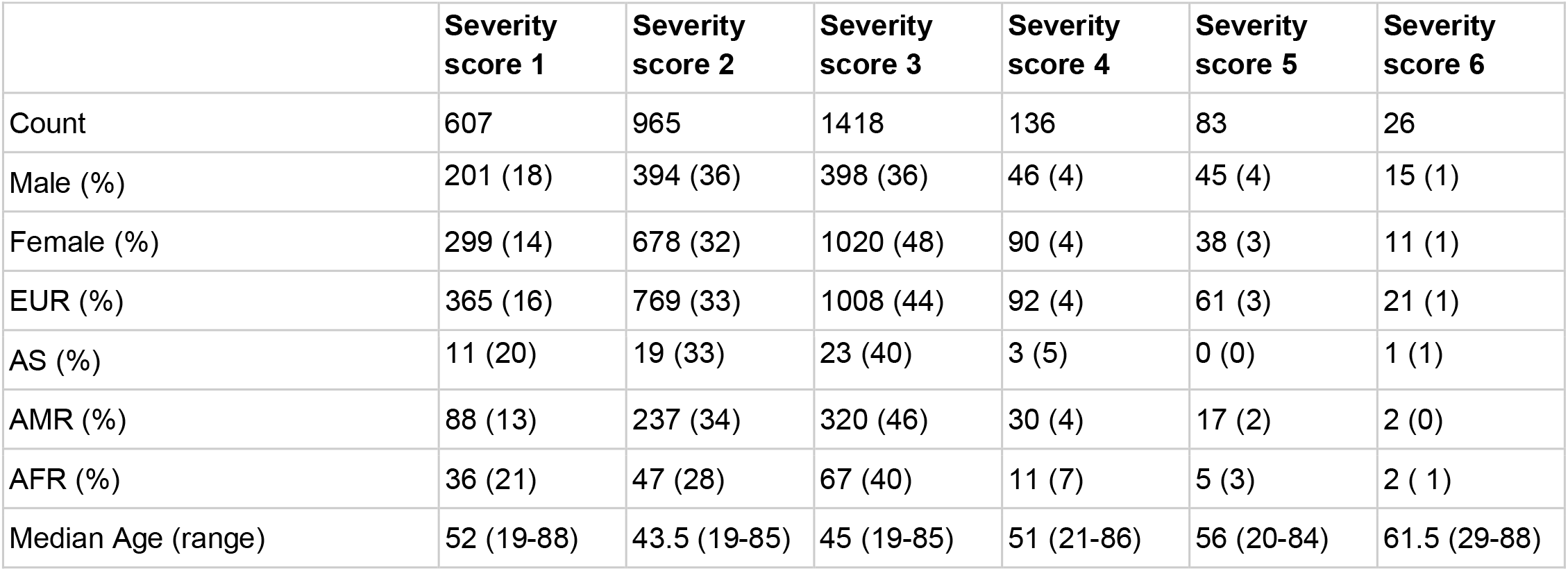
Demographic breakdown of the AncestryDNA cohort. Severity score 0 is left out since it corresponds to individuals who tested negative for SARS-CoV-2 infection. Severity scores 1-6 are described in Materials and Methods: Severity scoring and hospitalization.

|  | **Severity score 1** | **Severity score 2** | **Severity score 3** | **Severity score 4** | **Severity score 5** | **Severity score 6** |
| --- | --- | --- | --- | --- | --- | --- |
| Count | 607 | 965 | 1418 | 136 | 83 | 26 |
| Male (%) | 201 (18) | 394 (36) | 398 (36) | 46 (4) | 45 (4) | 15 (1) |
| Female (%) | 299 (14) | 678 (32) | 1020 (48) | 90 (4) | 38 (3) | 11 (1) |
| EUR (%) | 365 (16) | 769 (33) | 1008 (44) | 92 (4) | 61 (3) | 21 (1) |
| AS (%) | 11 (20) | 19 (33) | 23 (40) | 3 (5) | 0 (0) | 1 (1) |
| AMR (%) | 88 (13) | 237 (34) | 320 (46) | 30 (4) | 17 (2) | 2 (0) |
| AFR (%) | 36 (21) | 47 (28) | 67 (40) | 11 (7) | 5 (3) | 2 ( 1) |
| Median Age (range) | 52 (19-88) | 43.5 (19-85) | 45 (19-85) | 51 (21-86) | 56 (20-84) | 61.5 (29-88) |

We next assessed the contribution of genetic variability across the HLA locus to hospitalization as a binary outcome. Using BIGDAWG for case-control association analysis (20), we found no individual HLA types or specific amino acid variants across the HLA locus that were associated with hospitalization (Supplementary File 1). Only the DPB1 locus (p=0.04) was found to be associated with the binary outcome of whether an individual developed any COVID-19 symptoms.

To assess an individual’s capacity to present SARS-CoV-2 peptides, we computed HLA-specific MHC binding affinities of all k-mers of sizes between 8 and 12 inclusive from the SARS-CoV-2 proteome (n=48,395 unique peptides) passing a proteasomal cleavage propensity filter. We used two different predictive tools: netMHCpan and HLAthena (21,22). In agreement with our prior work (11), we find a wide variety in putative peptide presentation capacity across different HLA types (Supplementary Figure 1).

We next developed a pooled multivariate model of severity score, accounting for comorbidities as well as putative viral peptide presentation as a function of HLA type, and we found that age (p < 0.01), BMI (p < 0.01), asthma status (p < 0.01), diabetes status (p < 0.01), and other lung conditions (p < 0.05) were all predictive (Figure 1; Supplementary Table 1- Stratified_LM_models_corrected.csv). There was no association between the number of putatively presented class I peptides and COVID-19 severity. The significance of the association between BMI and age was diminished for age >60 years. This is consistent with CDC reports of obesity as a risk factor in hospitalization and death, specifically among individuals <65 years (3).

**Figure 1:**
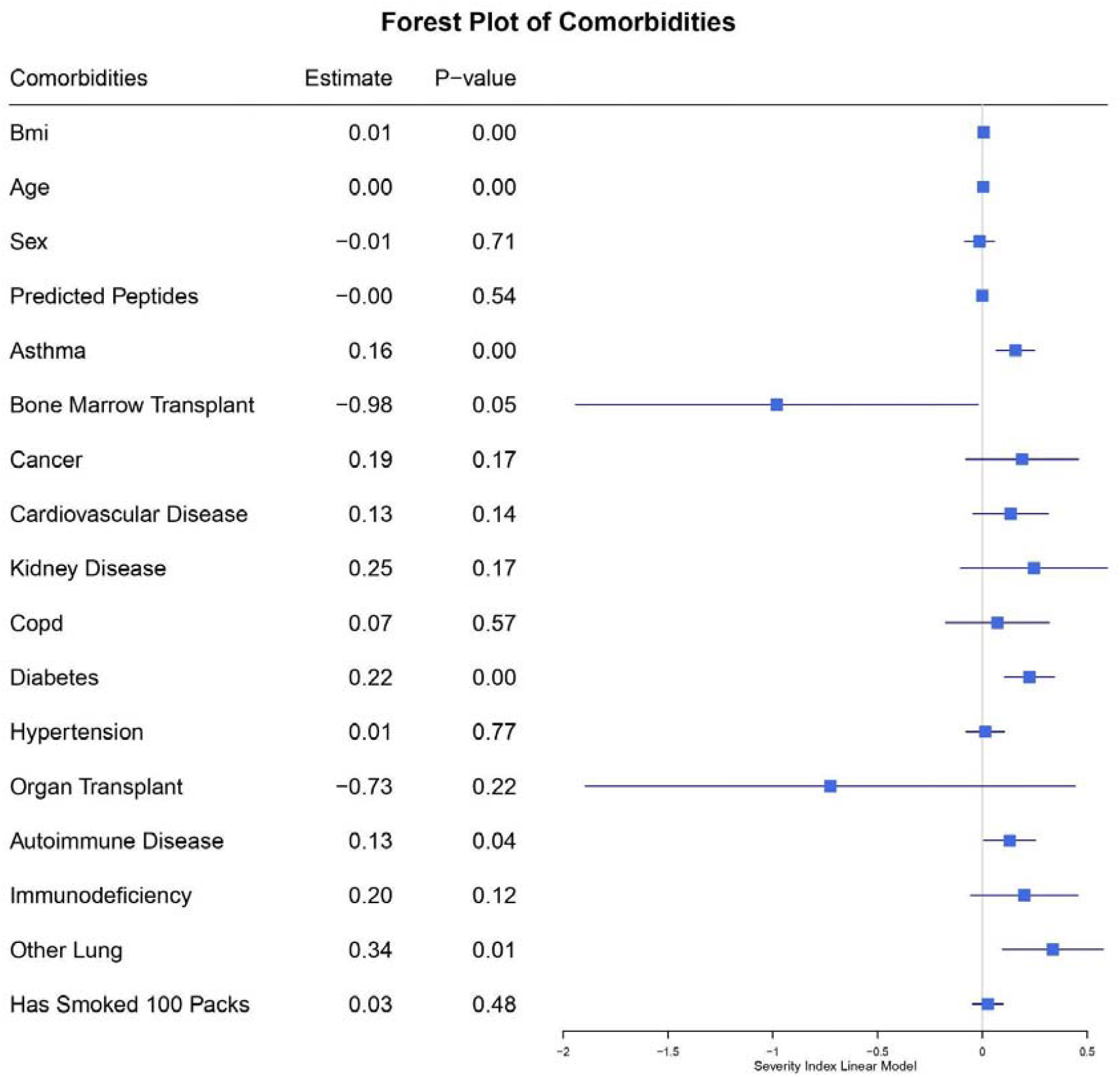
Forest plot of comorbidities including putative viral peptide presentation as a function of HLA type (Predicted Peptides) in the pooled multivariate model for predicting severity score. The estimate column shows the coefficients of variables in the linear model. Each line represents the 95% confidence interval for the estimate value. For the sex variable, female is 1 and male is 0. Positive values for the estimate are predicted to contribute to a higher severity score and vice versa for negative values.

**Figure 2:**
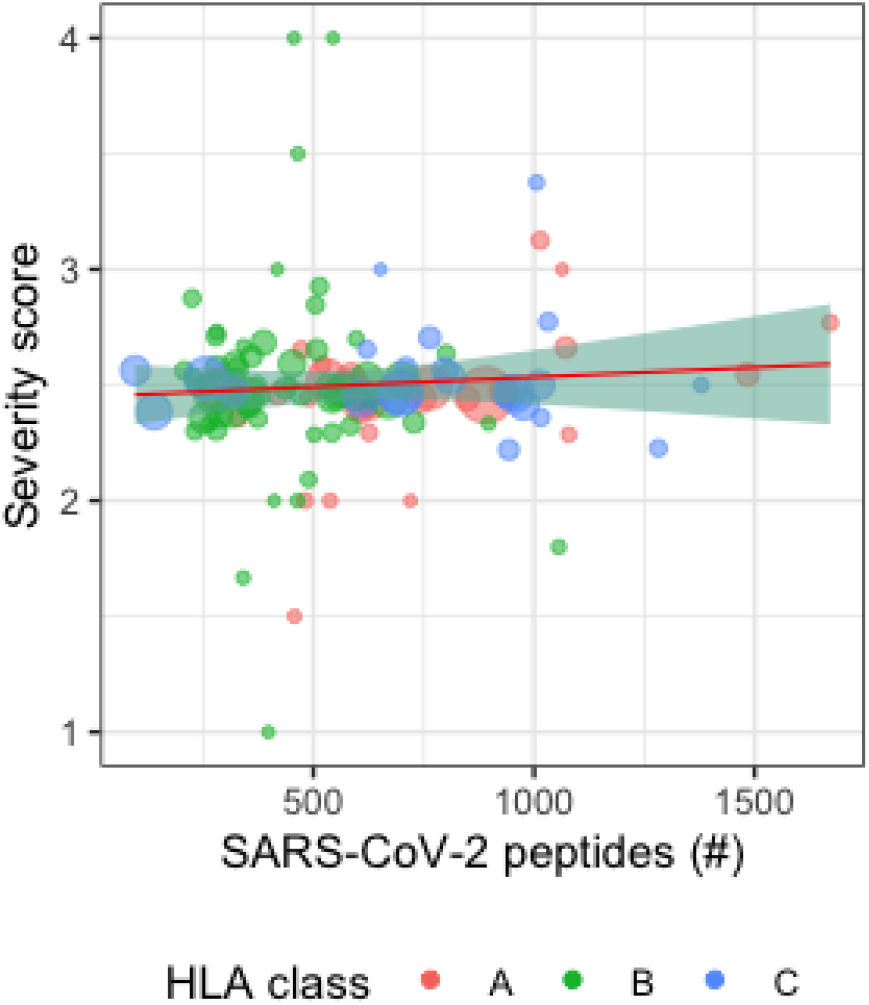
Scatter plot of HLA alleles with the number of predicted peptides vs. average severity score in the AncestryDNA dataset. Each data point represents a distinct HLA allele, with larger points representing larger numbers of individuals in the AncestryDNA dataset imputed to have the allele and the red, green, and blue colors representing HLA-A, HLA-B, and HLA-C respectively.

**Figure 3:**
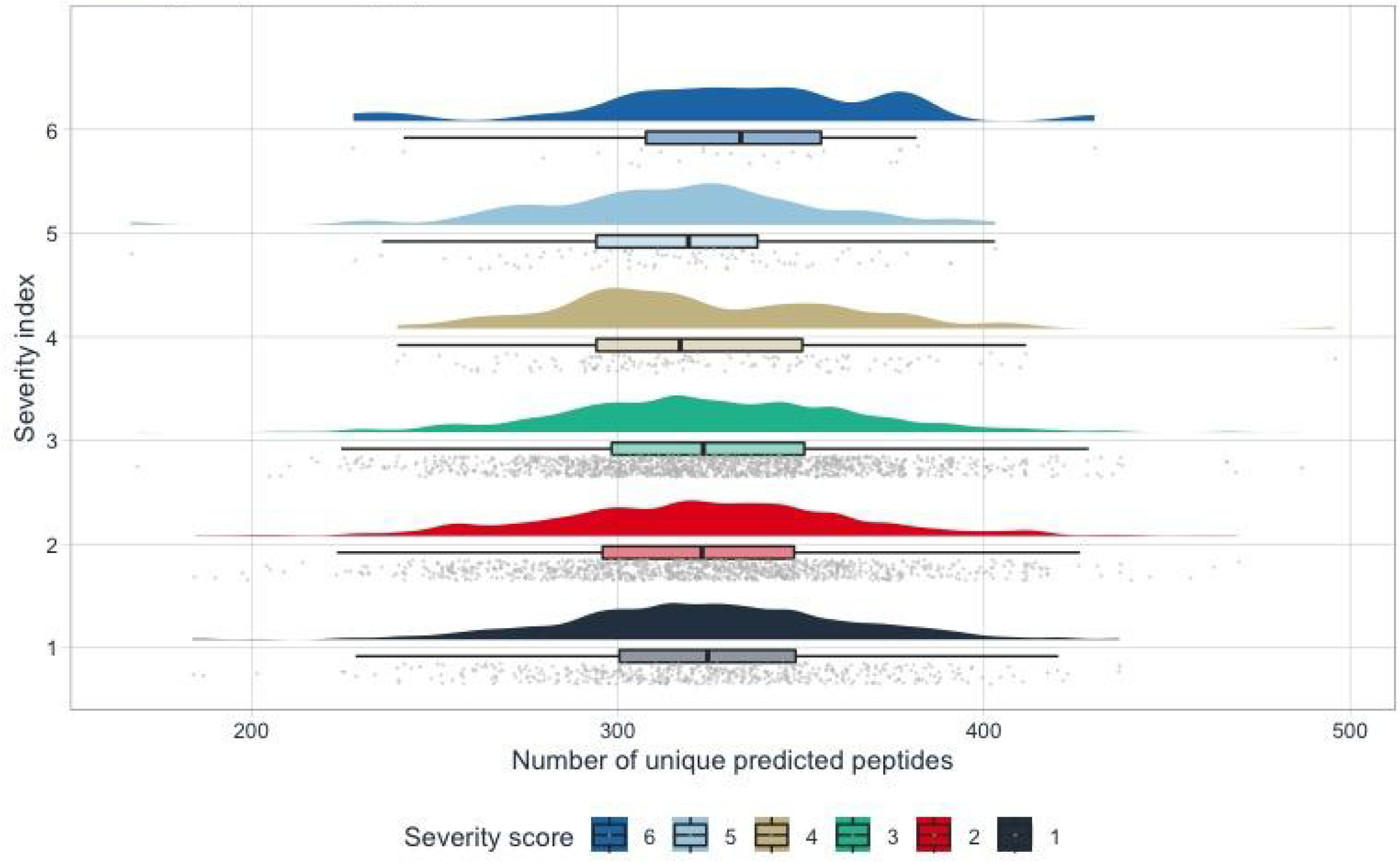
Distribution of unique predicted peptides vs. severity score. The half-violin plots represent the distribution of unique predicted peptides of the 3,235 AncestryDNA individuals who had the corresponding severity score. The boxplot shows the IQR of the unique predicted peptides and each point in the rain cloud below the boxplot represents the number of predicted peptides of each individual.

Predicted number of presented viral peptides demonstrated no significant relationship with disease severity when stratifying by individual ancestry, but the significance of various comorbidities was affected. Among individuals with EUR or AFR ancestry (2316 or 168 individuals, respectively) no clinical features were associated with disease severity score, while BMI (p<0.05), age (p<0.05), and hypertension (p<0.001) were all predictive of disease among individuals of AS ancestry (57 individuals), and BMI (p<0.05) was predictive among individuals of AMR ancestry (694 individuals).

## DISCUSSION

HLA genes are generally considered important for host response to novel infectious diseases. In this study, we found that age, BMI, and other comorbidities determined clinical outcome across 3,235 individuals as described in the literature (1–3,13), and to a far greater degree than an individual’s HLA-specific capacity to present SARS-CoV-2-specific peptides. While we previously explored the potential of HLA-peptide binding to predict COVID-19 severity (11), we do not see evidence for this phenomenon in the large real-world clinical cohort explored here. While the majority of the individuals were imputed to be of European ancestry, there were sizable numbers of individuals of Amerindian, Asian, and African descent. While Roberts et al. (10) performed a stratified GWAS analysis using this same dataset, with binary endpoints of hospitalization and whether an individual developed any COVID-19 symptoms, they did not specifically explore the role of HLA, which has a high level of variability that reduces power to detect differences in populations. Further, we investigated SARS-CoV-2 specific peptide presentation as a nonlinear function of HLA type, where some HLA types may be more similar to each other in the number of predicted peptides they can bind than they may be in canonical HLA supergroups.

We note several limitations to our work. Firstly, the proportion of SARS-CoV-2 peptides that we tested were generated through whole-peptidome *in silico* analysis of SARS-CoV-2. This may not be representative of the actual SARS-CoV-2 peptides presented in a given individual, whether due to biological sources such as viral variation, or methodological sources such as potential inaccuracies in peptide-MHC binding affinity predictions. Secondly, individuals who suffered debilitating infections may have been less likely to participate in the survey, and no individuals who died of COVID-19 were able to participate in the study, potentially resulting in an undercounting of the most severe phenotypes. Further, the cohort was primarily European, with much smaller sample sizes for African, Asian, and Amerindian ancestry. Lastly, these data were composed entirely of the unvaccinated cohort, as this population was tested and surveyed before the release of the many SARS-CoV-2 vaccines.

A number of other studies (15–18,23–25) have examined the relationship between HLA alleles and COVID-19 severity, and few have found alleles significantly associated with severity. In the majority of these studies, the large number of possible alleles in each study reduced the statistical power to identify significant alleles after multiple testing correction. Further, a number of studies reporting statistical significant associations between severity and HLA type were regional; they tended to have more ethnically and geographically homogeneous cohorts, likely resulting in overrepresentation of some alleles. Taken together with our analysis of the AncestryDNA dataset, we suggest that the literature does not reliably support the role of HLA type in modifying real-world COVID-19 disease severity across a population. There are multiple potential explanations for this, including that the data and analyses to date do not accurately reflect the true potential disease-modifying effects of HLA genes. On an individual basis, HLA type may indeed influence the severity of COVID-19 disease; however, this hypothesis is not readily borne out at a population level. Multiple demographic features and clinical comorbidities are significantly more predictive of disease severity in a population. Future work should take a very critical and individualized approach towards evaluating any connections between HLA variation and differences in COVID-19 disease severity.

## MATERIALS AND METHODS

### Ancestry imputation

Genetic ancestry was determined using plinkQC v1.9 (26) to combine genotypes of the cohort with genotypes of a reference dataset (27) consisting of individuals of known ethnicities. Principal component analysis (PCA) on the combined genotype panel was used to detect population structure of the reference dataset to the level of continental ancestry. A→T and C→G SNPs were removed from study and reference data as they are more difficult to align and only a subset of SNPs were required for the analysis. The study data was pruned for variants in linkage disequilibrium (LD) with an r^2^ > 0.2 in a 50kb window, and that list of pruned variants was used to reduce the size of the reference dataset. Checks were performed to ensure matching variant IDs and chromosomal positions between the study and reference dataset before merging and running PCA. Ancestries for the study population were then imputed from the principal components provided for the labeled reference dataset.

### HLA class I + II imputation

HLA Class I/II alleles were obtained using HIBAG v1.3 (28), a prediction method for HLA imputation that utilizes large training sets with known HLA and SNP genotypes in combination with attribute bagging. Ancestry-specific pre-fit models available within HIBAG for European, Asian, African, and Amerindian populations were applied to the subgroups of distinct ancestries within the AncestryDNA cohort.

### Severity scoring and hospitalization

We collapsed the 10 point WHO COVID-19 Ordinal Scale of disease severity (29) into a 7-point scale to accommodate available phenotype information in the AncestryDNA COVID-19 study.

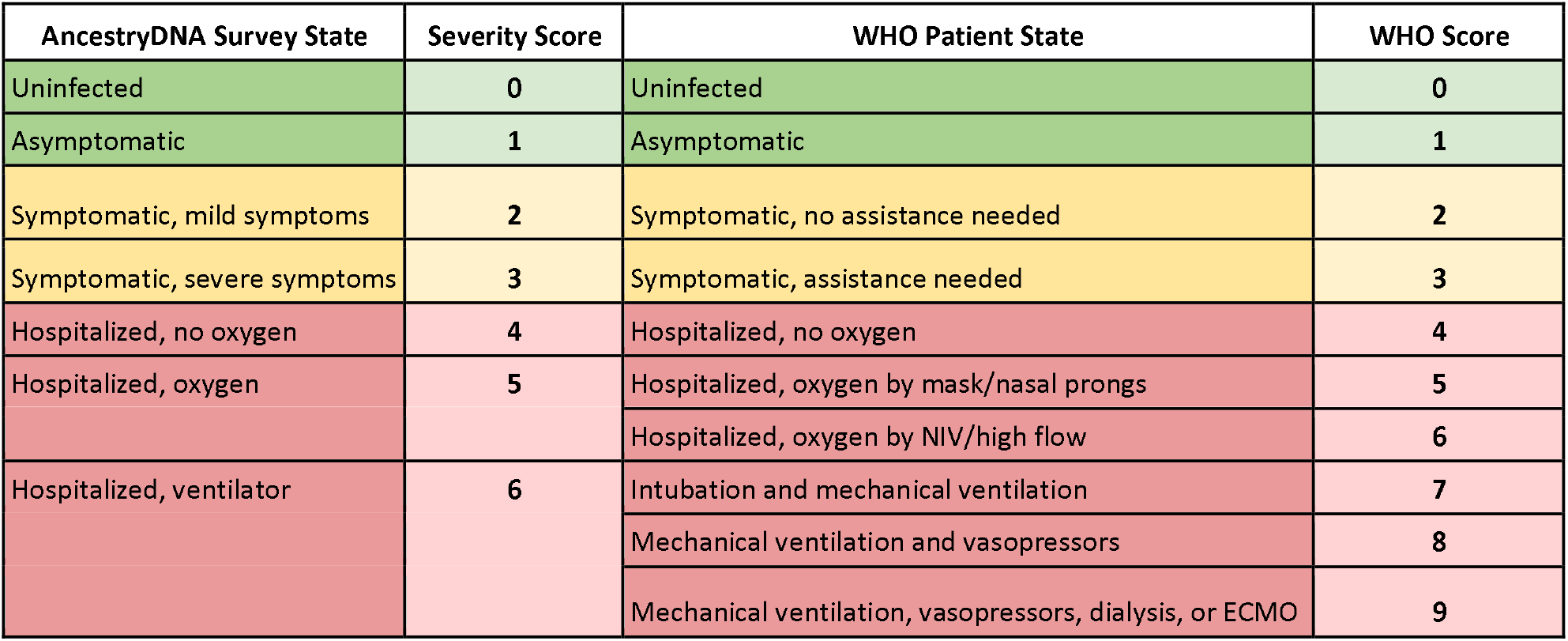

The possible symptoms in the AncestryDNA cohort are fever, shortness of breath, dry cough, body aches, abdominal pain, cough producing phlegm, and nausea. There are 3 levels of severity to each symptom: normal, severe, and very severe. We defined severe symptoms (Severity Score 3) as any of the listed symptoms at the severe or very severe level. In models where we used hospitalization as an endpoint, we added hospitalization as a binary variable, with scores >=4 considered hospitalized.

### HLA-peptide predicted binding

We obtained SARS-CoV-2 peptide sequences by k-merizing FASTA protein sequences obtained from the NCBI RefSeq database (NC_045512.2 and NC_004718.3) into 8- to 12-mers. These k-mers were filtered by NetChop v3.1 using default settings with a cutoff of 0.1. MHC class I binding affinity predictions were performed using netMHCpan v4.0 using the ‘-BA’ option to include binding affinity predictions and the ‘-l’ option to specify peptides 8 to 12 amino acids in length. Additional MHC class I binding affinity predictions were performed using HLAthena. For predicted peptide binding, we used the cutoff of <500nM for peptides predicted by netMHCpan v4.0 and the cutoff of >0.5 probability score for peptides predicted by HLAthena. While nearly all individuals have two HLA-A/B/C haplotypes constituting as few as three but as many as six distinct alleles, a single peptide may be predicted to bind to more than one of an individual’s HLA alleles. While there is no definitive evidence that a peptide is more likely to be presented when predicted to bind to more than one allele, we wanted to capture this possibility by using 2 metrics: an overall predicted peptide value and a unique predicted peptide value. For each individual, to calculate capacity to bind SARS-CoV-2 peptides, we summed the number of predicted peptides bound to each individual’s allele (min 3, max 6). For a unique-peptide specific capacity, the peptides were filtered to remove duplicates after summation.

### Statistical analyses

We performed statistical tests for HLA vs. hospitalization using the Bridging ImmunoGenomic Data-Analysis Workflow Gaps (BIGDAWG) pipeline and a comprehensive SARS-CoV-2 peptide-genotype binding analysis for all individuals in our dataset. All statistical analyses were performed using R version 4.0.3. For each statistical test, we performed pooled and ancestry-stratified testing. For multivariate linear modeling, we used the R function lm for multivariate regression with one of severity index, hospitalization status, or asymptomatic/symptomatic as the endpoint. Tests of Hardy-Weinberg equilibrium using Chi-squared testing for haplotypes, loci, and HLA-amino acid positions were performed using the BIGDAWG v1.3.4 R package. Note that all reported p-values have been corrected for multiple hypothesis testing, where relevant, using Benjamini-Hochberg correction.

## Data Availability

All data except restricted EGA data in the present study are available upon request to the authors.

https://ega-archive.org/studies/EGAS00001004716

