## Supplementary figures and images for "Minimal observed impact of HLA genotype on hospitalization and severity of SARS-CoV-2 infection"

### MS_nonbind_bind_plot.png

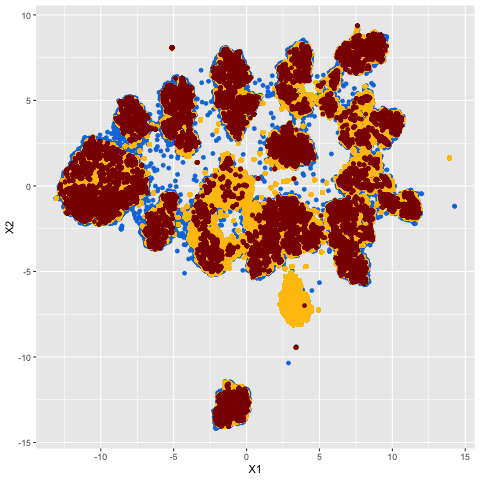

### non-binders_vs_massspec.png

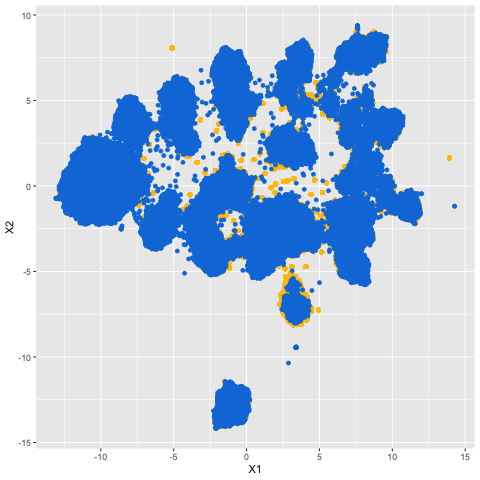

### Supplemental Figure 1

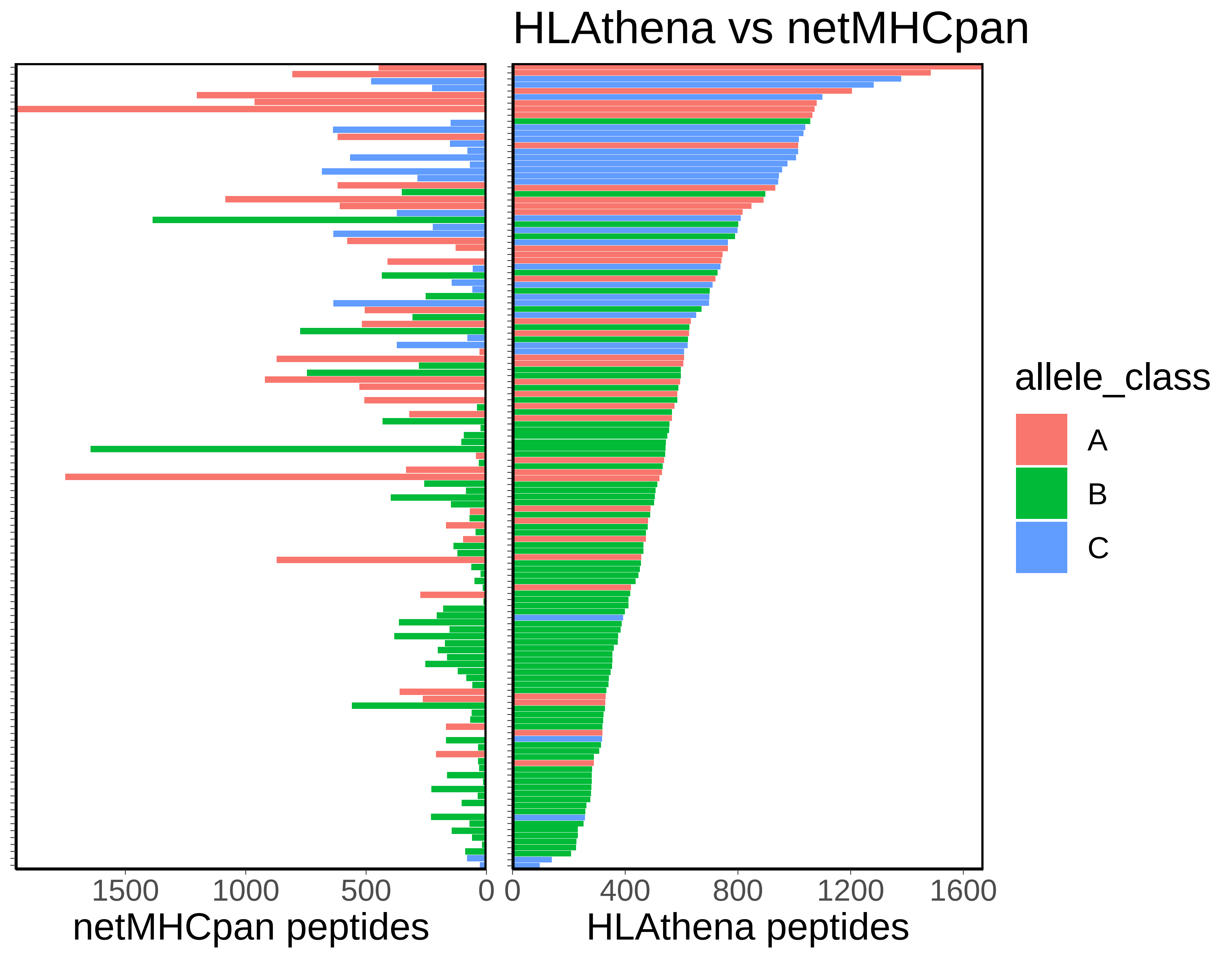
